# COVID-19 vaccination in pregnancy: views and vaccination uptake rates in pregnancy, a mixed methods analysis from the Born In Wales study

**DOI:** 10.1101/2022.05.09.22274769

**Authors:** M Mhereeg, H Jones, J Kennedy, M Seaborne, M Parker, N Kennedy, S Beeson, L Zuccolo, A Davies, S Brophy

## Abstract

**Background:** Vaccine hesitancy amongst pregnant women has been found to be a concern during past epidemics.

**Objectives:** The aims of this study were to 1) estimate COVID-19 vaccination rates among pregnant women in Wales and their association with age, ethnicity, and area of deprivation, using electronic health records (EHR) linkage, and 2) explore pregnant women’s views on receiving the COVID-19 vaccine during pregnancy using data from a survey recruiting via social media (Facebook, Twitter), through midwives, and posters in hospitals (Born in Wales Cohort).

**Design:** A mixed methods study utilising routinely collected linked data from the Secure Anonymised Information Linkage (SAIL) (Objective 1) and the Born In Wales Birth Cohort participants (Objective 2). SAIL combines data from general practice, hospital admissions, the national community child health dataset, maternal indicators dataset, and COVID-19 vaccination databases.

**Setting and participants:** Objective 1) All women documented as being pregnant on or after 13^th^ April 2021, aged 18 years or older, and eligible for COVID-19 vaccination were identified in routine health care. They were linked to the vaccination data up to and including 31^st^ December 2021. Objective 2) Separately, a cross-section of pregnant women in Wales were invited to complete an online survey via social media advertising. The survey asked what their views were on having the COVID-19 vaccination during pregnancy, and if they had already received, or intended to receive, the COVID-19 vaccination during their pregnancies. They were also asked to give reasons for their decisions.

**Outcomes:** 1 (a). Rate of vaccination uptake per month during pregnancy among women eligible for vaccination. 1 (b). Survival analysis was utilised to examine and compare the length of time to vaccination uptake in pregnancy, and variation in uptake by; age, ethnicity, and deprivation area was examined using hazard ratios (HR) from Cox regression. 2.Expectant mothers’ views of the COVID-19 vaccination during pregnancy.

**Results:** 

**Population-level data linkage (objective 1):** Within the population cohort, 32.7% (n = 8,203) were vaccinated (at least one dose of the vaccine) during pregnancy, 34.1% (n = 8,572) remained unvaccinated throughout follow-up period, and 33.2% (n = 8,336) received the vaccine postpartum. Younger women (<30 years) were less likely to have the vaccine and those living in areas of high deprivation were also less likely to have the vaccine (HR=0.88, 95% CI 0.82 to 0.95). Asian and other ethnic groups were 1.12 and 1.18 times more likely to have the vaccine in pregnancy compared to women of White ethnicity (HR=1.12, 95% CI 1.00 to 1.25) and (HR=1.18, 95% CI 1.03 to 1.37) respectively.

**Survey responses (objective 2):** 69% of participants stated that they would be happy to have the vaccine during pregnancy (n = 207). The remainder, 31%, indicated that they would not have the vaccine during pregnancy (n = 94). Reasons for having the vaccine related to protecting self and baby, perceived risk level, and receipt of sufficient evidence and advice. Reasons for vaccine refusal included lack of research about long-term outcomes for the baby, anxiety about vaccines, inconsistent advice/information, and preference to wait until after the pregnancy.

**Conclusion:** Potentially only 1 in 3 pregnant women would have the COVID-19 vaccine during pregnancy, even though 2 in 3 reported they would have the vaccination, thus it is critical to develop tailored strategies to increase its acceptance rate and to decrease vaccine hesitancy. A targeted approach to vaccinations may be required for groups such as younger people and those living in higher deprivation level areas.

## Introduction

Vaccination is acknowledged as a successful public health measure [1]. However, a growing number of the general population perceive vaccinations as unsafe and nonessential [1]. The SAGE working group described vaccine hesitancy using a ‘3 C’s’ model; Confidence, Complacency, and Convenience [2]. The model suggests that vaccine hesitancy arises when individuals a) do not have confidence in the vaccine’s safety and effectiveness b) do not believe in the seriousness of the disease and c) have the perception that access to the vaccine is inconvenient. Combatting the 3 C’s may lead to higher vaccine acceptance.

The issue of vaccine hesitancy was important during the COVID-19 pandemic. A literature review regarding vaccine hesitancy during the COVID-19 pandemic was conducted with fifteen studies being included in this review [3]. From this review, reasons for refusing the vaccine included being against vaccines in general, concerns about safety, thinking that a vaccine produced in a rush is too dangerous, general lack of trust, and doubts about the efficiency of the vaccine.

Vaccine hesitancy may be more prevalent in different populations [4]. For example, vaccine hesitancy may be more common in pregnant women [5]. During the COVID-19 pandemic, the limited data and change in advice/recommendations regarding the vaccination in pregnancy led to some hesitancy among pregnant women in certain settings [6]. Misleading information spread on social media platforms linking the COVID-19 vaccine to infertility reportedly lead to higher levels of distrust, and apprehension regarding the vaccine among pregnant women or those trying to conceive, as shown by studies from America [6].

In the UK, the COVID-19 vaccination programme started in December 2020 prioritising individuals at greater risk of being hospitalised or contracting severe cases of COVID-19 and individuals who care for vulnerable groups, such as health and social care workers [7]. At this time, the guidance from the UK’s Joint Committee on Vaccination and Immunisation (JCVI), was that the COVID-19 vaccine should not be given to pregnant women as there was a lack of data regarding the safety of COVID-19 vaccine during pregnancy. Later, in April 2021 the UK JCVI announced that pregnant women should be offered the COVID-19 vaccine [7].

Research is limited on population-level COVID-19 vaccine uptake in pregnancy in the UK. In Scotland, a national, prospective cohort study identifying ongoing pregnancies through extensive electronic health records (EHR) linkages showed vaccination rates in pregnant women to be substantially lower than in the general population; 32.3% in pregnant women compared to 77.4% in all women [8]. In England, 22.7% of women giving birth in August 2021 had received at least one dose of vaccine. This increased to 32.3% of women who gave birth in September - and the latest data shows that it rose to 53.7 in December 2021. Despite the overall increase in coverage, the uptake remains lower amongst pregnant women compared to the general population of the same age group [9]

Low vaccine uptake among pregnant women carries implications for both clinical and population health outcomes. Unvaccinated pregnant women are at increased risk of requiring hospital treatment for COVID-19 compared to those who are vaccinated [10]. Severe COVID-19 in pregnancy significantly increases the risks to the baby [11]. Pregnant women with severe COVID-19 were more likely to have a preterm birth, to have a pre-labour caesarean birth, to have a baby that was stillborn or to be admitted to a neonatal intensive care unit [11].

In research aimed to determine the attitudes towards vaccine acceptance and hesitancy of the COVID-19 vaccine in pregnant women [12], 300 pregnant women were surveyed using face-to-face methods, asking 40 questions about the COVID-19 pandemic and vaccination in January and February of 2021. It was observed that 37% of pregnant women (n = 111) stated their intention to receive the vaccine if it was recommended for pregnant women. The most common reasons stated for refusing the vaccine included lack of data about COVID-19 vaccine safety in pregnant populations and potential harm to the foetus. Pregnant women in the first trimester expressed higher acceptance of COVID-19 vaccination than those in the second and third trimesters. Therefore, this study reported low acceptance of the COVID-19 vaccination in a sample of pregnant women. Identifying attitudes towards the vaccine among pregnant women will be beneficial for generating vaccination strategies that increase uptake during the pandemic. However, the opinion on the vaccines may have changed over time.

The acceptance of the COVID-19 vaccine among pregnant women and mothers of young children was investigated in 16 countries across the world [13]. The strongest predictors of vaccine acceptance included confidence in vaccine safety or effectiveness, worrying about COVID-19, belief in the importance of vaccines to their own country, trust of public health agencies/health science, as well as attitudes towards routine vaccines [13].

The aims of this study are to Objective 1a) use national health data linkage of covid-19 vaccination and pregnancy records to identify vaccine uptake amongst pregnant women in Wales, Objective 1b) examining differences by age, ethnicity, and area of deprivation and Objective 2a) gain an insight into views and opinions on COVID-19 vaccine during pregnancy in a cross-section of pregnant women in Wales.

## Materials and methods

### Study design and setting

A cohort study utilising routinely collected linked data from the Secure Anonymised Information Linkage (SAIL) databank. Data sources include general practitioners (GP), hospital admissions, national community child health, maternal indicators, and vaccination databases. All women recorded as being pregnant on or after 13^th^ April 2021, aged 18 years or older, and eligible for COVID-19 vaccination were identified. They were linked to the COVID-19 vaccination dataset for dates up to and including 31^st^ December 2021.

Pregnant women in Wales were invited through the Born in Wales study to complete an online survey via social media (Facebook, Twitter), recruitment through midwives, and posters in hospitals. Respondents were either pregnant or gave birth during the COVID-19 pandemic between the 1^st^ November 2021 to the 24^th^ March 2022 when the questionnaire was live. The main open ended questions employed were ‘what is your view on having the COVID-19 vaccine in pregnancy?’, and ‘have you had, or would you have, the COVID-19 vaccine while pregnant and why?’. All responses were anonymous, the self assessed inclusion criteria was living in Wales and either being pregnant or having had a baby during the pandemic.

### Data sources and linkage

Analysis was undertaken using individual level linked routinely collected national-scale data available in the SAIL databank [14,15], which anonymously links a wide range of person-based data using a unique personal identifier. The linkage is brought together under the Born In Wales study [16] and includes GP records linked with hospital admission (inpatient and outpatient) records, the National Community Child Health (NCCHD), Maternal Indicators (MIDS) and the Vaccination data. The GP system utilises READ codes, which are 5-digit codes that relate to diagnosis, medication, and process of care codes. The secondary care dataset uses ICD-10 codes for diagnosis and surgical interventions. The NCCHD comprises of information pertaining to birth registration, monitoring of child health examinations, and immunisations. The MIDS dataset contains data relating to the woman at initial assessment and to mother and baby (or babies) for all births. In addition to these datasets, the Welsh Demographic Service dataset was linked to extract information on deprivation. In particular, the Welsh Index for Multiple Deprivation (WIMD) quintile was employed as a proxy to assess social deprivation. These records were linked at the individual level for all women known to be pregnant in Wales between 13^th^ April 2021 and 31^st^ December 2021 and then stratified for subanalysis by age group, ethnicity, and WIMD quintile. Quality of linkage has been assessed and reported as 99.9% for GP records and 99.3% for hospital records [17]. All linkage was at the person level.

### Ethical approval

The data held in the SAIL (Secure Anonymised Information Linkage) databank in Wales/UK are anonymized. All data contained in SAIL has the permission from the relevant Caldicott Guardian or Data Protection Officer and SAIL-related projects are required to obtain Information Governance Review Panel (IGRP) approvall. The IGRP approval number for this study is 0911. The Research Ethics Service approval was also given by the North West-Greater Manchester East Research Ethics Committee for the qualitative research. The Ethics Reference Number is RIO 030-20.

### Study population and key dates

Pregnant women were identified as any woman who had pregnancy codes in the GP data, in hospital admissions for pregnancy, or mothers in the NCCHD or MIDS databases with the baby birth date (Pregnancy end date) and gestational age at birth available. The baby’s birth date and gestational age enabled the start date of pregnancy to be determined for those who gave birth during the study period. Data collected included the vaccination data, Welsh index of multiple deprivation (WIMD), and ethnicity. The WIMD is an official measure for the relative deprivation of areas of Wales. It combines eight separate domains of deprivation, each compiled from a range of different indicators (income, employment, health, education, access to services, housing, community safety, and physical environment) into a single score and is widely used to measure deprivation in Wales.

As for the general population, pregnant women were first offered the vaccine in December 2020 if they were (1) health or care workers, as these positions have increased risk of SARS-Cov-2 infection or (2) in a high risk group due to health conditions. Since April 2021, pregnant women have been offered the vaccine as part of the standard age based rollout of the vaccination programme [18]. We selected the 13^th^ April 2021 as the study start date because phase 2 of the vaccination program, which aimed to provide vaccinations to individuals aged 40 to 49, 30 to 39, and 18 to 29 years, started on this date. The inclusion criteria were pregnant women who had not received the vaccination or had one dose of vaccination before pregnancy, alive, known pregnant on the first day of follow up, and aged 18 years or older. The exclusion criteria were women who were fully vaccinated (i.e two vaccinations) before pregnancy, or those for whom it was not possible to determine the start date of pregnancy due to unavailability of the gestational age and initial assessment dates in their records. Women were censored at the birth, death, or moved out of Wales while pregnant.

### Calculating pregnancy start date

Pregnancy start dates were calculated from the following sources:

For pregnancies identified from the NCCHD and MIDS datasets the pregnancy start dates were calculated based on the gestational age and the week of birth data items available in these databases. In cases where gestational age is missing, a value of 40 weeks was used as the majority of with missing data (92%) had birth weights suggestive of full term infants. Thus, the pregnancy start date (last menstrual period) was simply calculated by subtracting the gestational age at birth (in weeks) from the week of birth. Pregnancies identified from both datasets were compared/matched and duplicate records were removed.

For pregnancies identified from the GP dataset, all pregnant women with a pregnancy code and event date that occurred during the study period were extracted (Supplementary table 1). For those identified from the hospital admissions data (PEDW), all women with a pregnancy diagnosis code and an attendance date occurring during the study period were also extracted (Supplementary table 2). Identified cases from both the GP and PEDW were separately matched to those identified from the NCCHD and MIDS datasets to include only those who are still pregnant. Furthermore, the identified cases from both resources were further matched for removing duplicates, and then linked to the initial assessment-related data items in the MIDS dataset. The gestational age in weeks and initial assessment data items are available in order to calculate the pregnancy start date. In cases where multiple records were found per pregnant woman, only the first occurring record between the study dates of interest was selected. The pregnancy start date for every successfully linked case was then calculated by subtracting the gestational age from the initial assessment date.

### Statistical analysis

Descriptive statistics were conducted on rates of vaccination uptake per month during pregnancy among women eligible for vaccination, stratified by age group. Uptake rates were also gathered on ethnicity, and area of deprivation stratified by age group. Kaplan-Meier survival analysis was employed to examine time to vaccination and censored at the birth, death, or moved out of Wales while pregnant. The log rank test was used to determine if there were differences in the survival distributions of vaccine uptake times within the different demographic variables. Differences were reported in median times (MD) with 95% confidence intervals and significance level accepted at p<0.05. Multivariate Cox regression hazard models were utilised to examine the impact of the explanatory variables age group, ethnicity, and area of deprivation jointly on vaccination uptake, reporting hazard ratios (HR) with 95% confidence intervals and significance level accepted at p<0.05. The reference groups were those aged 25-29, white ethnicity, and those living in the most affluent area. The data handling and preparation for the descriptive statistics, survival analysis and Cox proportional hazard modelling were performed in an SQL database within SAIL databank utilising Eclipse. Final data preparation specific to these analyses such as setting the reference groups was performed in IBM SPSS Statistics 28. Descriptive statistics were performed in Microsoft Excel 2016 and Survival/Cox regression analyses were performed in SPSS.

### Survey Methods

Expectant mothers or mothers who gave birth during the COVID-19 pandemic were invited to complete an online survey via social media advertising. Codebook thematic analysis [19] was used to generate themes from an open-ended question on the survey: ‘What is your view on having the COVID vaccination in pregnancy, have you or would you have the COVID vaccination when pregnant and why?’. Thematic analysis identifies and describes patterns across data [19]. Analysis involved six phases 1) data familiarisation and writing familiarisation notes 2) systematic data coding 3) generating initial themes from coded and collated data 4) developing and reviewing themes 5) refining, defining, and naming themes and 6) writing the report. All data were independently analysed by HJ and SB, who then discussed their findings. This was to ensure that important concepts within the data were not missed, and to achieve a richer understanding of the data through multiple perspectives.

## Results

A total of 28,343 women who were known to be pregnant on 13/04/2021 or became pregnant after this date were identified. Excluding those who were fully vaccinated before pregnancy (n = 3,232), the cohort comprised 25,111 pregnant women. Those women were followed up and their records were linked to the COVID-19 vaccination data up to and including 31/12/2021. (Figure 1 describes the participants in the cohort). Most of the women were aged between 30-39, and between 25-29 years (48.4% and 29.7% respectively). The majority were White (77.8%). Nearly a quarter were living in the most deprived quintile (23.3%) and 14.4% were in the least deprived quintile (Table 1).

**Fig. 1.**
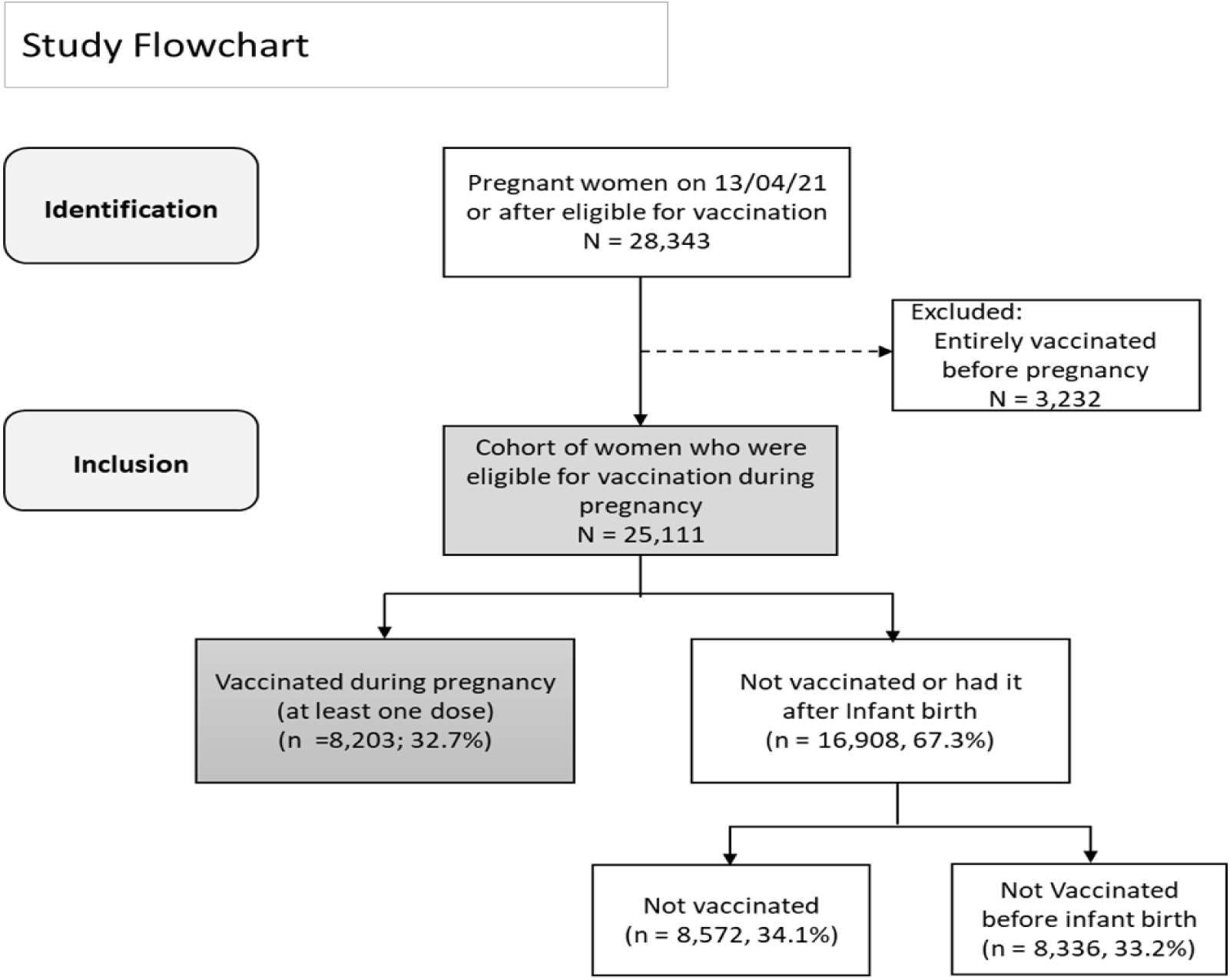
Flowchart of the cohort identification.

**Table 1.** Descriptive summaries of the pregnant women eligible for vaccination

|  |  | N | % |
| --- | --- | --- | --- |
| Age | 18-24 | 4,664 | 18.6% |
|  | 25-29 | 7,447 | 29.7% |
|  | 30-39 | 12,143 | 48.4% |
|  | 40-50 | 857 | 3.4% |
| **Ethnicity** | White | 19,547 | 77.8% |
|  | Asian | 902 | 3.6% |
|  | Other | 571 | 2.3% |
|  | Mixed | 316 | 1.3% |
|  | Black | 440 | 1.8% |
|  | Unknown | 3,335 | 13.3% |
| **WIMD Quintile** | 1<sup>st</sup> (Most deprived) | 5840 | 23.3% |
|  | 2<sup>nd</sup> | 4795 | 19.1% |
|  | 3<sup>rd</sup> | 4157 | 16.6% |
|  | 4<sup>th</sup> | 3804 | 15.1% |
|  | 5<sup>th</sup> (Least deprived) | 3626 | 14.4% |
|  | Unknown | 2889 | 11.5% |

### Uptake of COVID-19 vaccination in pregnancy

Over the study window, only 32.7% (n = 8,203) of pregnant women received the vaccine (i.e had at least one dose of the vaccine) during pregnancy, 34.1% (n = 8,572) were not vaccinated, and 33.2% (n = 8,336) had the vaccine after the birth. From the start of the vaccination programme on the 7^th^ of December 2020, there was a slow growth in the uptake of the vaccine among pregnant women as only those who were health or care workers or in the at risk group were first offered the vaccine. Uptake of the vaccine rose rapidly in April 2021 when pregnant women were offered the vaccine as part of the standard age-based rollout of the vaccination program; 32.7% of pregnant women were vaccinated by the end of December 2021 (Figure 2a). The vaccine uptake each month was consistently lower in younger women (<30 years) compared to those aged 30 or older. Overall, only 23.5% of those aged 18-24 were vaccinated by the end of December 2021 compared to 40.3% in those aged 40-50 (Figure 2b, Supplementary table 3). Starting from April, vaccine uptake rates started rising rapidly among those aged 40-50 with 21.7% of them receiving the vaccine, followed by those aged 25-29 and 30-39 rising rapidly in May (31.7% and 32.4% respectively), and then in June for those aged 18-24 (23.1%). Uptake rates were slower thereafter for all groups (Figure 2c, Supplementary table 3).

**Fig. 2a.**
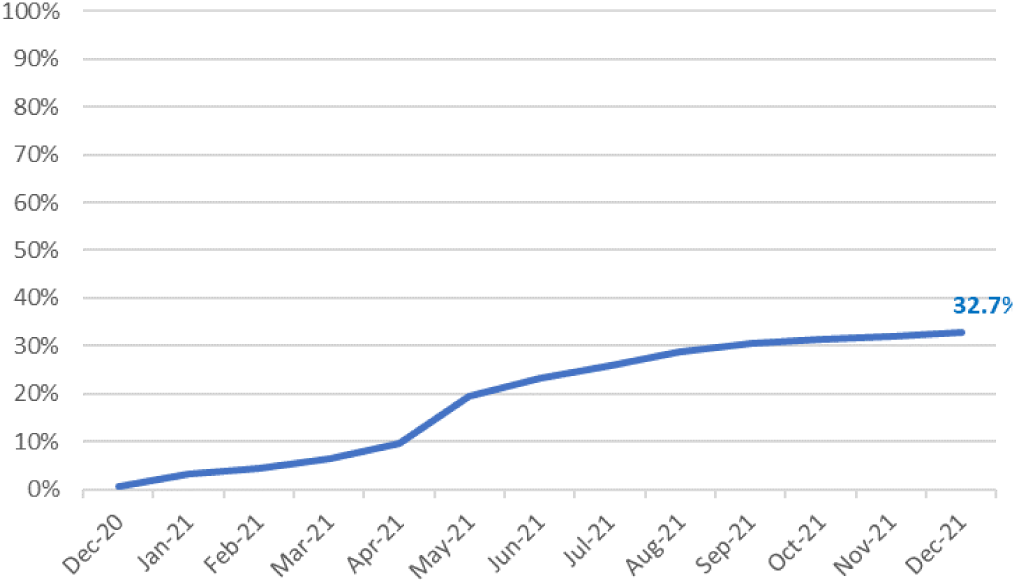
Cumulative vaccine rates by month.

**Fig 2b.**
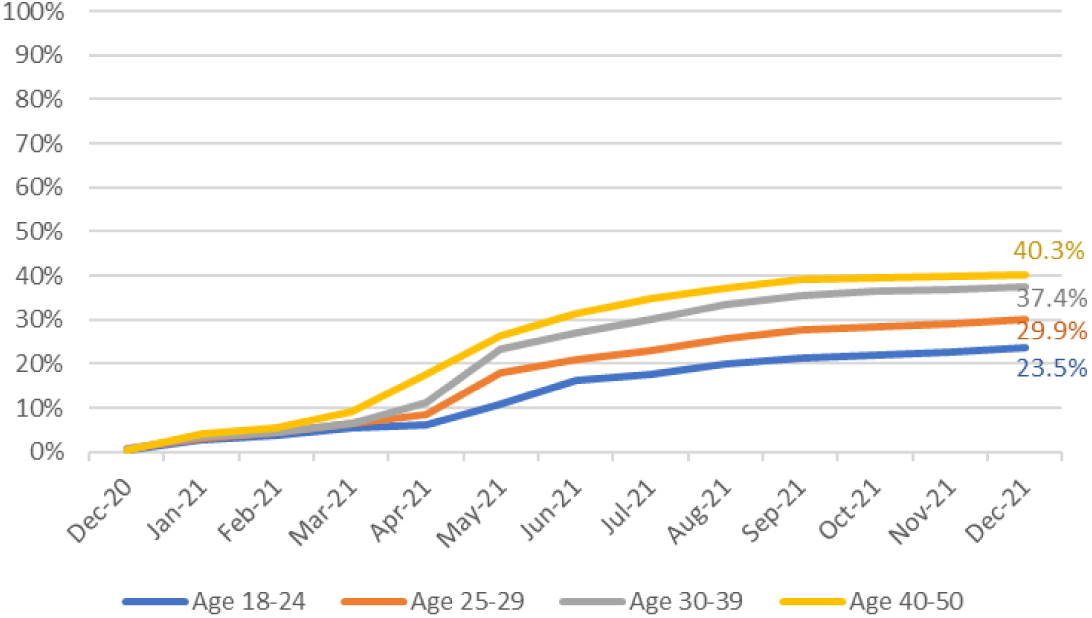
Cumulative vaccine rates by month and age group.

**Fig 2c.**
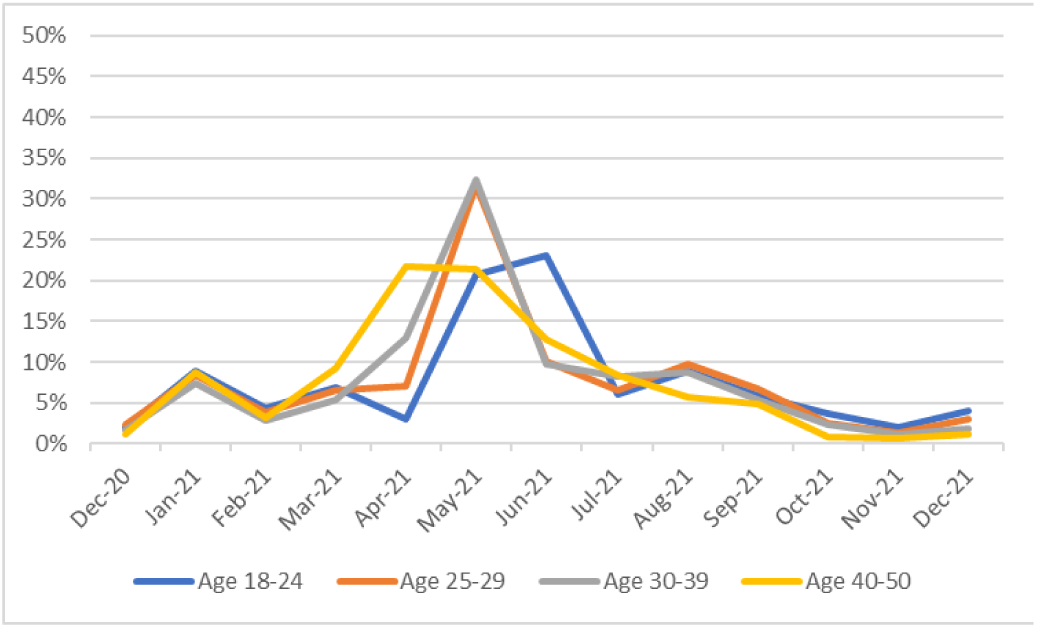
Vaccine uptake rates by month and age group.

The uptake rate was higher among Asian women (36.7%, 95% CI 33.6 to 39.8) compared to White (33.9%, 95% CI 33.2 to 34.5) women and the other ethnic groups (34.0%, 95% CI 30.1 to 37.9), especially compared to women of Mixed (23.7%, 95% CI 19.0 to 28.4) or Black ethnicity (23.9%, 95% CI 19.9 to 27.8), where less than a quarter of women had the vaccine (Figure 3a). The uptake for those aged 18-24 in the Black ethnic group was 14.7% lower than those aged 18-24 in the Asian ethnic group, and 25% lower compared to their peers aged 40-50. Figure 3b shows that the uptake was highest among Asian women and lowest among Black and Mixed groups for all age groups. The uptake rate for those living in the most deprived area is 16.4% lower than those living in the least deprived area. The biggest difference is in those aged 30 or older. In the 30-39 and 40-50 age groups there are 14.9% and 17.8% difference between the most and least deprived areas despite the fact that uptake in general is higher in those groups (Figures 3c, 3d, Supplementary table 4).

**Fig. 3a.**
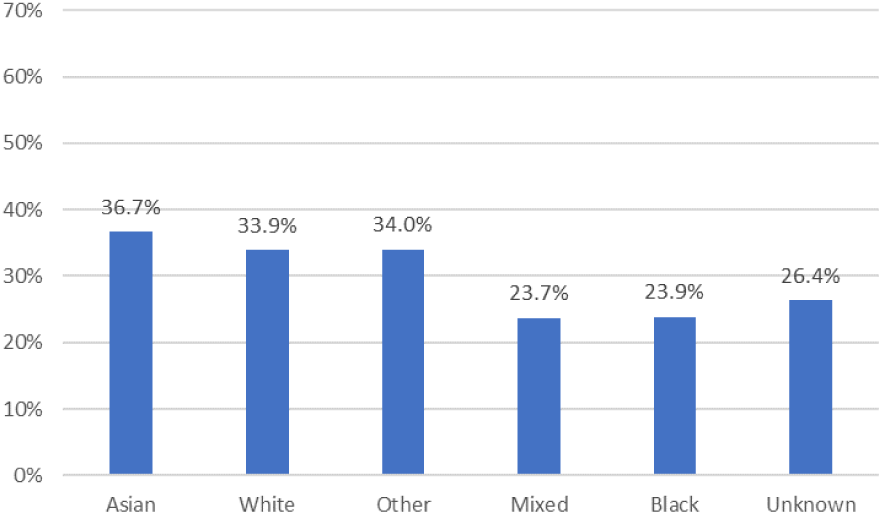
Vaccine uptake rates by ethnicity.

**Fig 3b.**
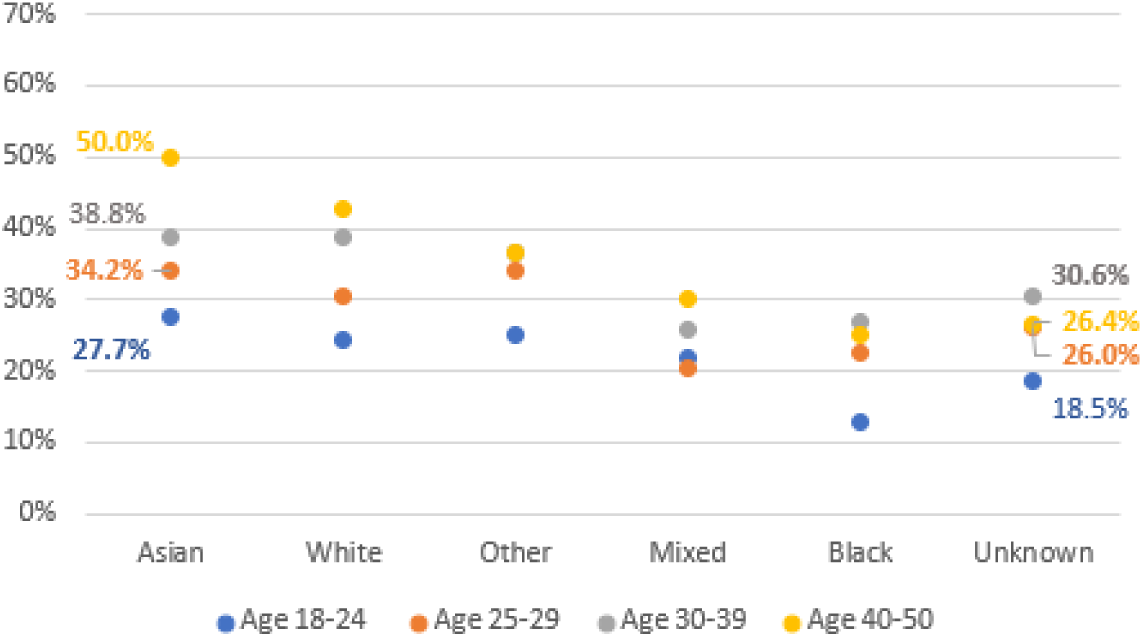
Vaccine uptake rates by ethnicity and age group.

**Fig. 3c.**
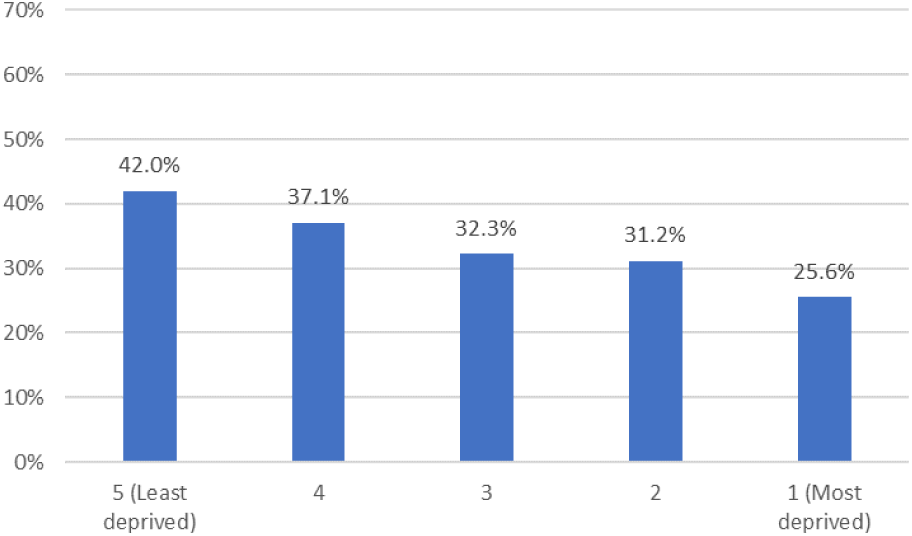
Vaccine uptake rates by WIMD quintile area of deprivation.

**Fig. 3d.**
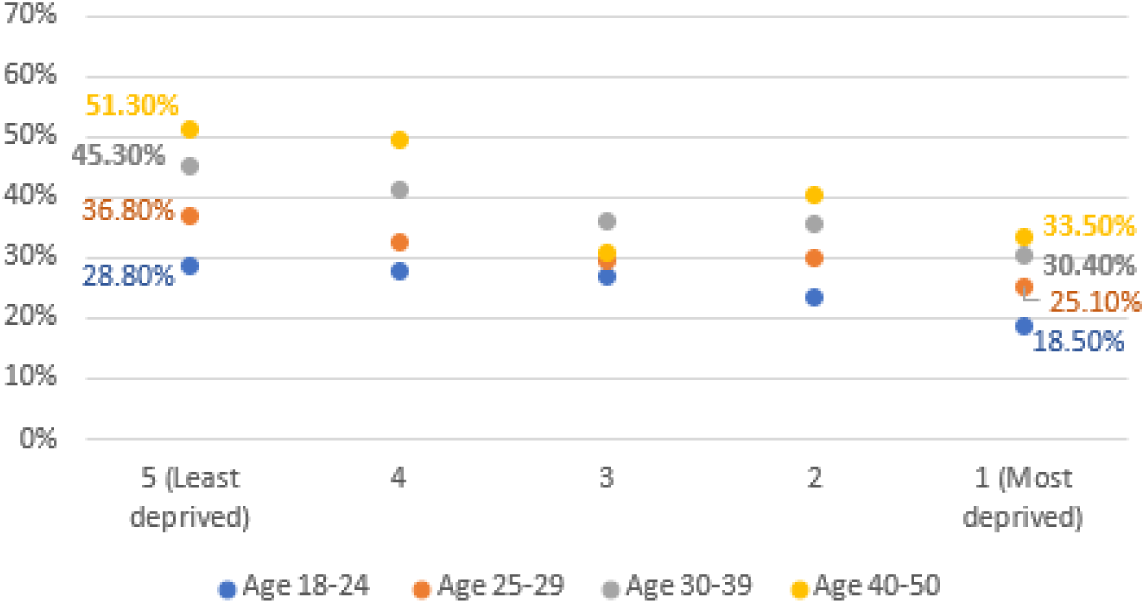
Vaccine uptake rates by WIMD quintile area of deprivation and age group.

### Examining time to first vaccination in pregnancy

Kaplan-Meier survival analysis shows that women aged 18-14 and 25-29 had identical median times to vaccine uptake of 136 days (95% CI 128.5 to 143.5), and 136 days (95% CI 131.6 to 140.4) respectively. This was longer than those aged 30-39 or 40-50, which had median times to vaccine uptake of 115 days (95% CI 111.6 to 118.4) and 99 days (95% CI 87 to 111) respectively. A log rank test was conducted to determine if there were differences in the survival distributions of vaccine uptake times for the different groups. The survival distributions were statistically significantly different, X^2^(3) = 72.5, p< .001. Pairwise log rank comparisons were conducted to determine which groups had different survival distributions. There were statistically significant differences between those aged 18-24 vs. those aged 30-39, X^2^(1) = 30.3, p<.001, and 18-24 vs. 40-50 groups, X^2^(1) = 26.7, p<.001. The same is mirrored in age group 25-29 vs age groups 30-39 and 40-50. However, the survival distributions for groups 18-24 vs. 25-29 and 30-39 vs. 40-50 were not significantly different (Fig 4a, Supplementary table 5).

**Fig 4a.**
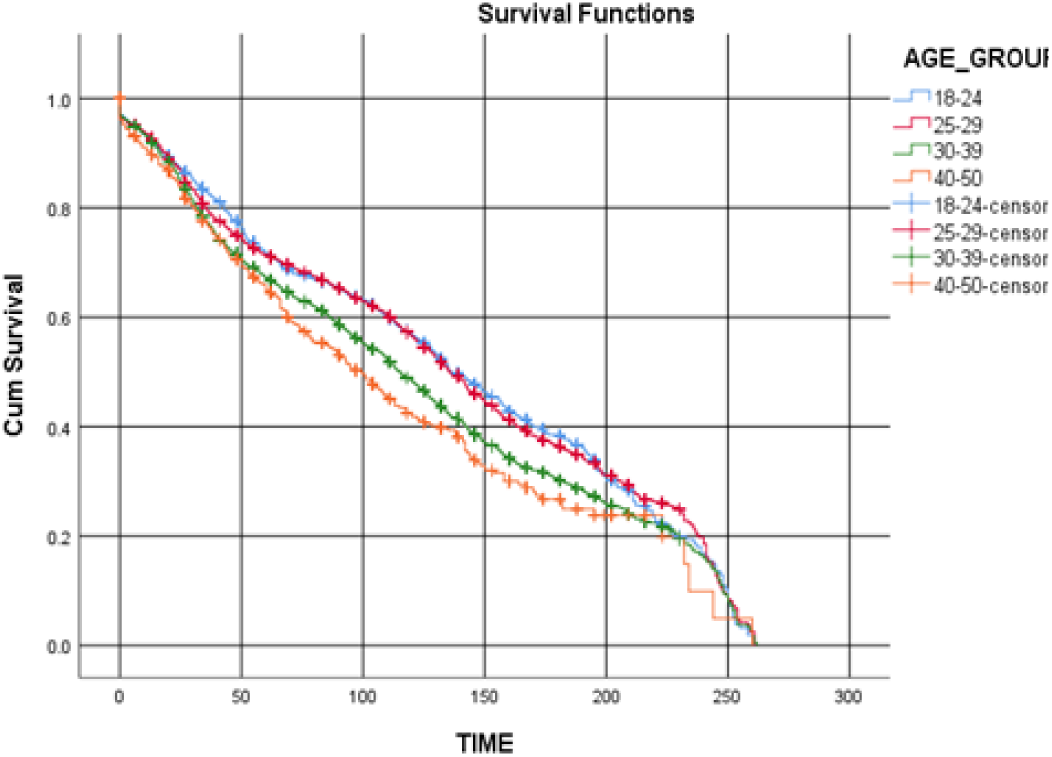
Time to vaccine uptake in pregnancy by age.

**Fig 4b.**
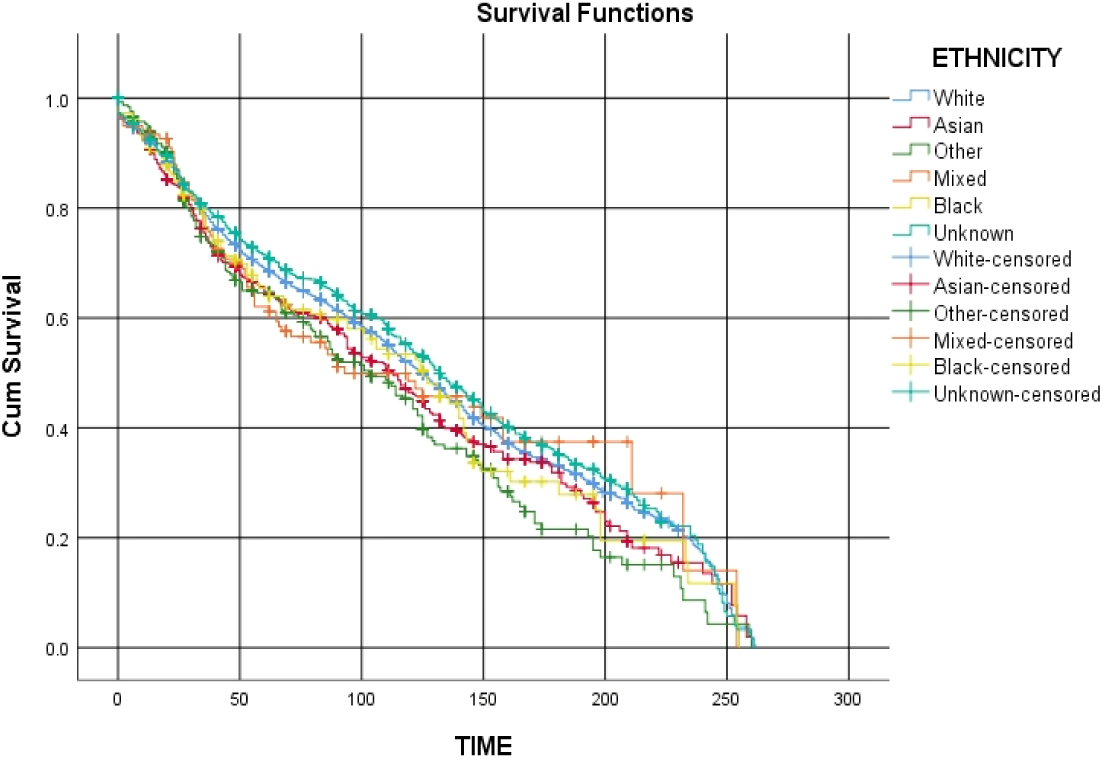
Time to vaccine uptake in pregnancy by ethnicity.

**Fig 4c.**
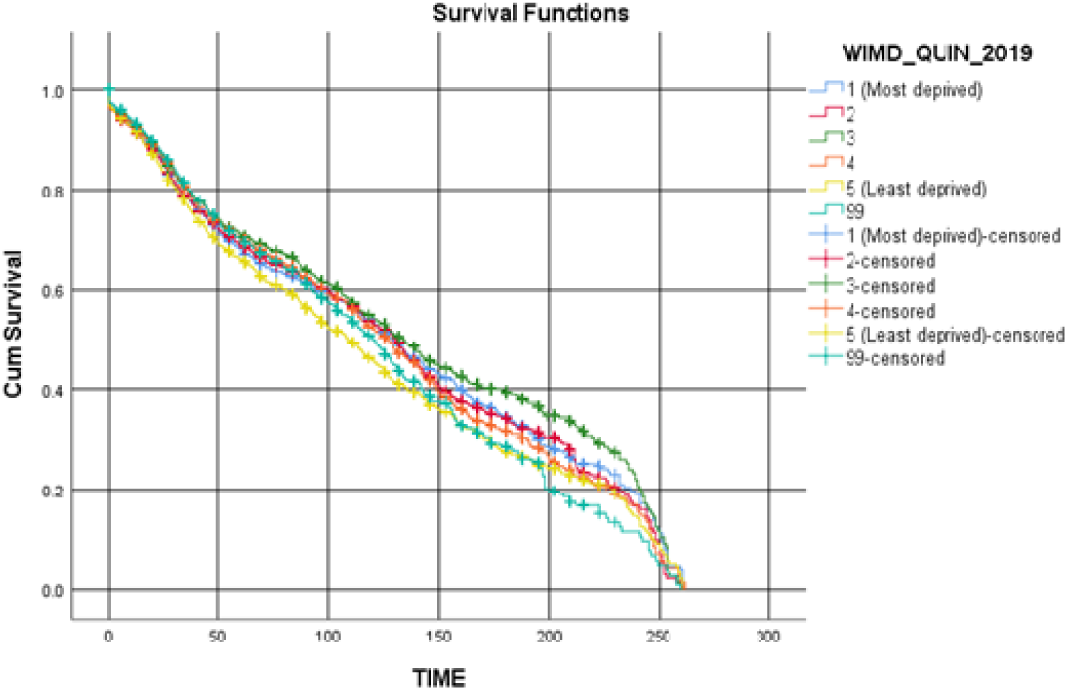
Time to vaccine uptake in pregnancy by WIMD.

The survival distributions between certain ethnic groups were significantly different, X^2^(5) = 16.7, p=0.005. The Asian and Other ethnic groups had median times of 113 days (95% CI 100.3 to 125.7), and 103 days (95% CI 85.4 to 120.6), which were less than the White’s median time of 125 days (95% CI 122.4 to 127.6). These differences were significant between the Asian vs. the White groups, X^2^(1) = 4.2, p=0.04, and the Other vs. the White group, X^2^(1)=6.4, p=0.01 (Fig. 4b, Supplementary table 5). Those who are living in the most deprived area had a median time to vaccine uptake of 129 days (95% CI 123 to 135.1). This was longer than those living in the least deprived area, which had the lowest median time of 109 days (95% CI 103.2 to, 114.8). The survival distributions between the different deprivation levels were significantly different, X^2^(5) = 41.9, p<.001. There were significant differences between those living in the least deprived areas and those living in the most deprived area X^2^(1)=17.5, p<.001, and all the other areas of deprivation (p<.001 for all) (Fig. 4c, Supplementary table 5).

### Examining the impact of age, ethnicity and deprivation area on vaccine uptake

Multivariate Cox regression was conducted to examine the variations in uptake by age group, ethnicity, and area of deprivation jointly on vaccination acceptance. Those aged 40-50 were 1.33 times more likely to have the vaccine compared to those aged 25-29 (HR=1.33, 95% CI 1.18 to 1.49, p<.001), also those aged 30-39 were 1.17 times more likely to have the vaccine compared to those aged 25-29 (HR=1.17, 95% CI 1.11 to 1.23, p<.001) (Table 2). The Asian and Other (the majority of other were Chinese) ethnic groups were 1.12 and 1.18 times more likely to have the vaccine compared to the White group (HR=1.12, 95% CI 1.00 to 1.25, p=.047) and (HR=1.18, 95% CI 1.03 to 1.37, p=.021) respectively. It was also observed that the vaccine uptake was lower among those living in the most deprived areas compared to those living in the most affluent areas (HR=.88, 95% CI 0.82 to 0.95, p<.001).

**Table 2.** Cox Regression analysis of factors associated with vaccination uptake among pregnant women eligible for vaccination, adjusted analysis. HR - hazard ratio. CI - confidence interval.

| Characteristic |  | HR (95% CI) | P value |
| --- | --- | --- | --- |
| **Age** | 25-29 | Reference |  |
|  | 18-24 | .99 (0.92 – 1.07) | .796 |
|  | 30-39 | 1.17 (1.11 – 1.23) | <.001 |
|  | 40-50 | 1.33 (1.18 – 1.49) | <.001 |
| **Ethnicity** | White | Reference |  |
|  | Asian | 1.12 (1.00 – 1.25) | .047 |
|  | Other | 1.18 (1.03 – 1.37) | .021 |
|  | Mixed | 1.02 (.81 – 1.29) | .855 |
|  | Black | 1.08 (.89 – 1.31) | .440 |
|  | Unknown | .93 (.87 – 1.003) | .060 |
| **WIMD quintile** | 5<sup>th</sup> (Least deprived) | Reference |  |
|  | 4<sup>th</sup> | .90 (.83 – .96) | .003 |
|  | 3<sup>rd</sup> | .81 (.76 – .88) | <.001 |
|  | 2<sup>nd</sup> | .91 (0.84 – 0.98) | .008 |
|  | 1<sup>st</sup> (Most deprived) | .88 (0.82 – 0.95) | <.001 |

### Mothers views of COVID-19 vaccination in pregnancy

There were 331 women who had a baby during the pandemic or who were currently pregnant between the 1st November 2021 and 24th March 2022 and participating in Born in Wales. 44.4% of the women were aged between 30-39 and the majority were White (82.2%) (Supplemantry table 6). They answered the open question ‘What is your view on having the COVID-19 vaccination in pregnancy, have you or would you have the COVID-19 vaccination when pregnant and why?’ 68% of women said they would be happy to have the vaccine in pregnancy (n = 224) and 32% said they would not have the vaccine in pregnancy (n = 107). Two main themes of ‘Happy to have the vaccine’ and ‘Concerns about the vaccine’ were generated (Table 3).

**Table 3.**
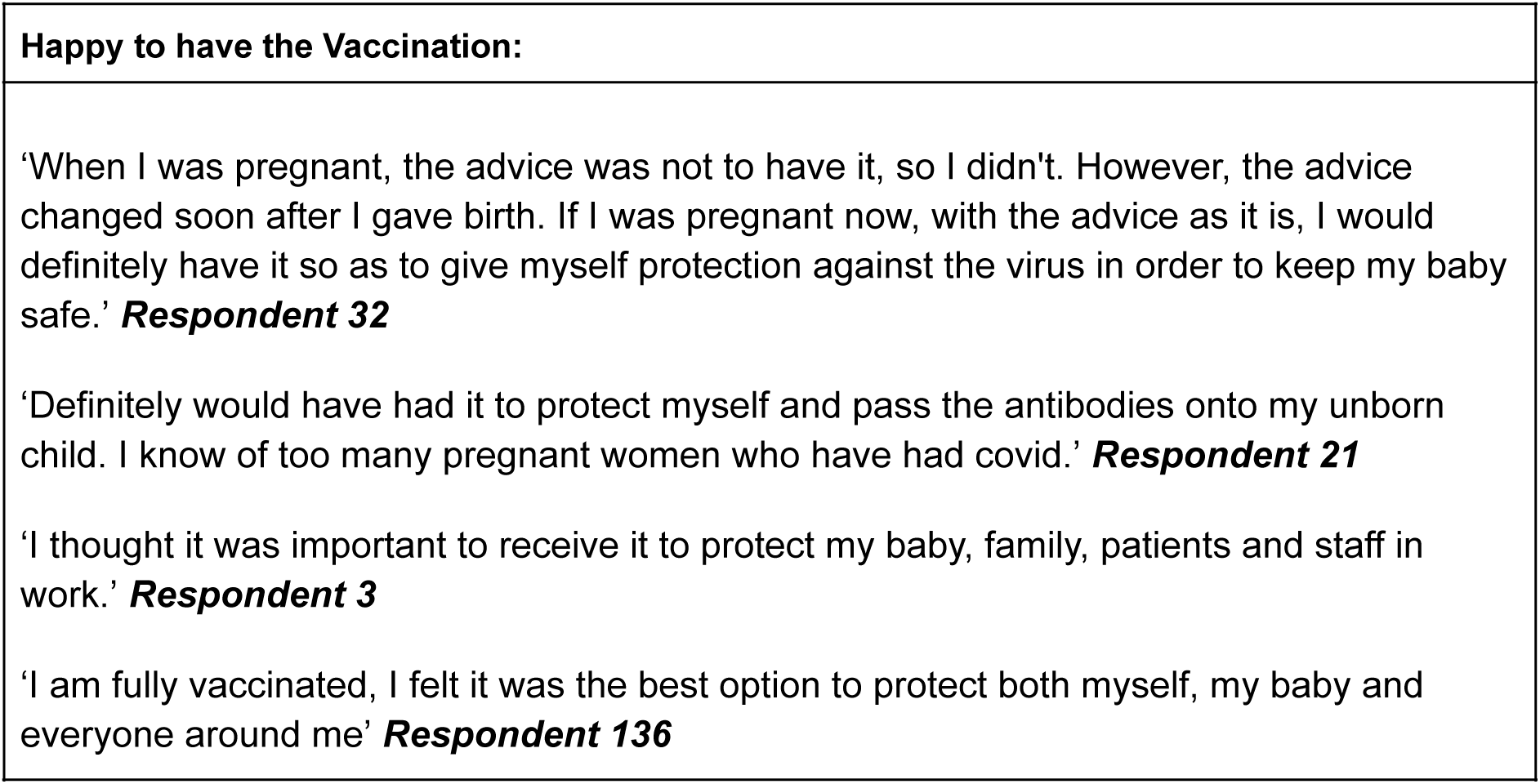
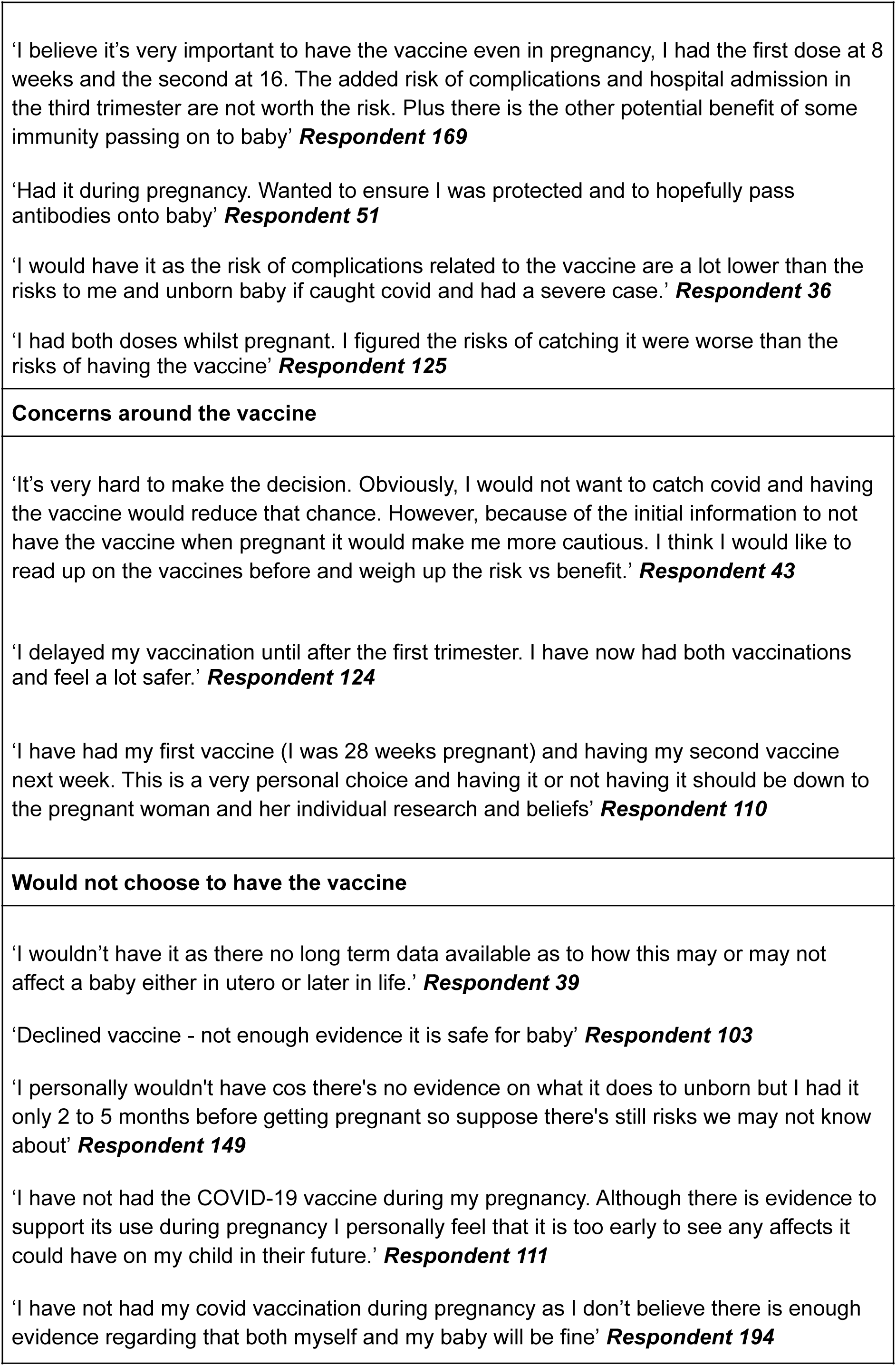
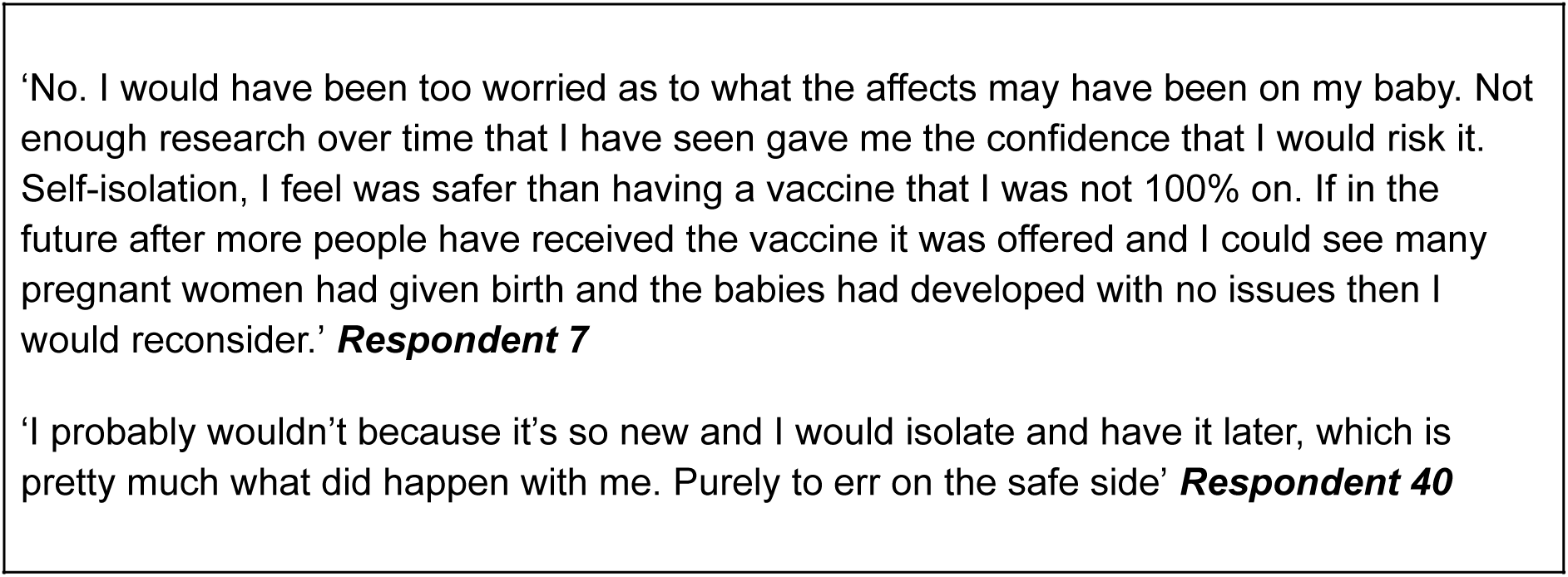
Themes emerging from responses to the question ‘What is your view on having the COVID-19 vaccination in pregnancy, have you or would you have the COVID-19 vaccination when pregnant and why?’

Those who were happy to have the vaccine felt it offered protection for their unborn baby and themselves, they felt it might help pass antibodies onto their unborn child and felt the chance of complications and hospital admission were not worth the risk. However, some women were more cautious as guidelines had changed and said they would want to read up more to understand the risk and benefits. Others felt that it was a very personal choice and should be up to the pregnant women and that it was difficult as there was a lot of misinformation and changes in advice was confusing. Those who would not be happy to have the vaccine predominately felt there was not enough long-term data available especially regarding babies’ safety. They felt self isolation was better protection and a number of women felt it was better to wait until after the birth.

## Discussion

This study describes the uptake rates of the COVID-19 vaccination and reasons for vaccine hesitancy or vaccine acceptance in pregnant women in Wales. From the linked data, 34.1% of pregnant women chose not to have the vaccine, 32.7% of the cohort received the vaccine in pregnancy and 33.2% had the vaccine after their baby was born. These findings reflect what was observed in qualitative responses where 31% of pregnant women responding stated that they would not have the COVID-19 vaccine during pregnancy. Our findings are similar to the overall high vaccine rates in the UK population where as of March 2022, 77% of the UK population has had at least one dose of the COVID-19 vaccine [20]. Across the world there is variation in vaccine uptake with 96% of the population fully vaccinated in the United Arab Emirates, 78% in France and 66% in the US [20].

Our qualitative results highlighted reasons for hesitancy including concerns over long term safety to the baby and confusion regarding changing recommendations, these findings are in step with previous vaccine hesitancy research studies [3,6]. However there are changing attitudes over time. For example, a literature review conducted in 2020 indicated that there were high levels of uncertainty regarding the vaccine [3], which may highlight higher levels of vaccine hesitancy compared to now as more research has been conducted regarding the safety of the vaccine. It also included reasons why some women were not hesitant and were pro-vaccination which could potentially inform how to address the hesitancy of others.

From the linked data, age, ethnicity, and deprivation level appeared to influence whether expectant mothers chose to have the vaccine or not and this re?ects patterns of uptake in the general population. The youngest age group (age 18-24) were least likely to have the vaccine and the oldest group (age 40+) were most likely to have the vaccine. Research has found that being younger is associated with both refusal and delay of the COVID-19 vaccine in

Portugal [21]. Moreover, studies have indicated evidence of reduced vaccine uptake in younger women aged <30 who gave birth in London between March 1, 2020, and July 4, 2021 [22]. Vaccine hesitancy was also higher in younger age groups (26.5% in 16-24 year olds compared to 4.5% in those aged 75+) [23]. Vaccine uptake was substantially lower in pregnant women in Scotland than in the general female population; 32.3% pregnant women compared to 77.4% in all women [8].

The rate of vaccine uptake differed significantly between certain ethnic groups. Asian and Other (e.g. Chinese ethnicities predominantly) were most likely to have the vaccine and differed significantly from those of White ethnicity. Research has found that one of the highest acceptance rates was observed in China, with an average of 77.4% of women accepting a future vaccine during pregnancy [23] which may explain our findings of higher vaccine acceptance in asian women. In the Black and mixed ethnicity groups, vaccination rates were the lowest. Willingness to be vaccinated was generally high across the UK population [24]. However, vaccine hesitancy does exist in population subgroups [24]. Black and Pakistani/Bangladeshi ethnic groups had higher levels of vaccine hesitancy from responses to a survey [24].

This research showed those living in the least deprived areas in Wales were more likely to have the vaccine compared to those living in the most deprived areas. The characteristics of recipients of the COVID-19 vaccine in England have also been investigated [25]. Research found that there were differences in vaccination uptake in various subgroups including ethnic groups (White 42.5% vaccinated, Black 20.5% vaccinated) and deprivation level (least deprived 44.7% vaccinated, most deprived 37.9% vaccinated) [25]. Similarly, there was evidence of reduced vaccine uptake in younger pregnant women with high levels of deprivation in the UK [22].

### Strengths and limitations

The study has several strengths, it utilises primary and secondary health care data for pregnant women in Wales including the maternity and child health data, it gives a national perspective of COVID-19 vaccine hesitancy, making the findings generalisable due to its total population cohort. Our qualitative survey questions allowed a free text response asking participants to provide their opinion on the vaccine and any reasons why they would or would not have it. These responses gave a true insight into the thoughts and feelings of pregnant women in Wales during the pandemic. Findings showing that the reasons for not wanting a vaccine included anxiety about the vaccine, change in advice and information or prefer to delay until after the birth.

The mixed methods design used in this study provided rich detailed information firstly about population-level vaccination uptake rates as well as rich qualitative responses from a cross-section of pregnant women in Wales. Using the two methods provides us with more insight into the reasons why 34.1% of pregnant women refused the vaccine completely and may inform vaccine strategies moving forward.

Vaccinations protect against severe disease. As the pandemic continues, booster vaccinations are increasingly important to provide protection against severe COVID-19, especially in vulnerable populations such as pregnant women. However, the data shows that 67.3% of pregnant women did not receive the vaccine in pregnancy. The world health organisation (WHO) recommends the COVID-19 vaccination in pregnant women when the benefits of vaccination to the pregnant woman outweigh the potential risks [26]. The priority should be to vaccinate pregnant women and encourage their vaccination.

The study had some limitations in that the qualitative analysis did not indicate in which trimester pregnant women had the vaccine as it has been reported that pregnant women in the first trimester expressed higher acceptance of COVID-19 vaccination than those in the second and third trimesters [12]. From our qualitative responses, expectant mothers expressed that they wanted to wait until later in their pregnancies before accepting the vaccine. Some commented that they would even wait until after the birth. The preference of accepting the vaccine after birth was reflected in the quantitative analysis, where 33.2% of pregnant women had the vaccine after childbirth.

## Conclusion

In conclusion, it is critical to develop tailored strategies to increase the acceptance rates of the COVID-19 vaccine and decrease hesitancy. A more targeted approach to vaccinations may need to be addressed to reach certain groups such as younger people, black and mixed ethnic minorities, and those living in more deprived areas. Encouraging vulnerable populations including pregnant women is a priority moving forward.

## Data Availability

All data produced in the present study are available upon reasonable request to the authors

## Acknowledgements

This study is part of the National Centre for Population Health and Wellbeing, which is funded by Health Care Research Wales. This study makes use of anonymised data held in the Secure Anonymised Information Linkage (SAIL) Databank (15, 27, 28). We would like to acknowledge all the data providers who make anonymised data available for research.

This work was supported by Health Data Research UK, which receives its funding from HDR UK Ltd (HDR-9006) funded by the UK Medical Research Council, Engineering and Physical Sciences Research Council, Economic and Social Research Council, Department of Health and Social Care (England), Chief Scientist Office of the Scottish Government Health and Social Care Directorates, Health and Social Care Research and Development Division (Welsh Government), Public Health Agency (Northern Ireland), British Heart Foundation (BHF) and the Wellcome Trust.

This work was additionally supported by funding from the Data and Connectivity National Core Study, led by Health Data Research UK in partnership with the Office for National Statistics and funded by UK Research and Innovation(grant ref MC_PC_20058), with additional support by The Alan Turing Institute via ‘Towards Turing 2.0’ EPSRC Grant Funding.

The responsibility for the interpretation of the information supplied is the authors’ alone.

## Contributors

SB, MM, HJ, JK conceived the article. MM carried out the quantitative analysis. HJ and SB carried out the qualitative analysis. MM and HJ prepared the first draft of the manuscript. All authors made intellectually important contributions to the manuscript, wrote or edited parts of it, and approved the final version submitted for publication.

