## Supplemental Tables for "COVID-19 vaccination in pregnancy: views and vaccination uptake rates in pregnancy, a mixed methods analysis from the Born In Wales study"

Supplementary table 1: Read codes (v2) used to identify pregnancy from the primary care data (GP)

| Read code | Description |
|---|---|
| 13H7. | Unwanted pregnancy |
| 2711. | O/E - fundus 12-16 week size |
| 2712. | O/E - fundus 16-20 week size |
| 2713. | O/E - fundus 20-24 week size |
| 2714. | O/E - fundus 24-28 week size |
| 2715. | O/E - fundus 28-32 week size |
| 2717. | O/E - fundus 34-36 week size |
| 4453. | Serum pregnancy test positive |
| 4654. | Urine pregnancy test positive |
| 584.. | Ultrasound in obstetric diagnosis |
| 5841. | U-S obstetric scan requested |
| 5842. | U-S obstetric scan normal |
| 584B. | Viability US scan |
| 584C. | Antenatal ultrasound result received |
| 584D. | Antenatal ultrasound confirms intra-uterine pregnancy |
| 584Z. | U-S obstetric diagnosis scan NOS |
| 62... | Patient pregnant |
| 621.. | Patient currently pregnant |
| 6212. | Pregnant - blood test confirms |
| 6214. | Pregnant - on history |
| 6216. | Pregnant - planned |
| 6217. | Pregnant - unplanned - wanted |
| 621C. | Unplanned pregnancy |
| 621Z. | Patient pregnant NOS |
| 622.. | Antenatal care: gravida No. |
| 623.. | A/N care: obstetric risk |
| 625Z. | A/N care: social risk NOS |
| 628Z. | A/N risk NOS |
| 62A.. | A/N care provider |
| 62A3. | A/N - shared care |
| 62B.. | Delivery booking place |
| 62B3. | G.P. unit delivery booking |
| 62B4. | Consultant unit booking |
| 62B8. | Midwife unit delivery booking |
| 62C.. | Delivery booking - length of stay |
| 62F.. | Antenatal amniocentesis |
| 62G.. | Antenatal ultrasound scan |
| 62GB. | Antenatal ultrasounds scan at 4-8 weeks |
| 62GZ. | Antenatal ultrasound scan NOS |

| | |
|---|---|
| 62L.. | Antenatal blood group screen |
| 62N.. | Antenatal examinations |
| 62N1. | A/N booking examination |
| 62N3. | A/N 16-week examination |
| 62O1. | Fetal movements felt |
| 62X.. | Length of gestation |
| 62Y.. | Routine antenatal care |
| 62a.. | Pregnancy review |
| 62b.. | Antenatal HIV screening |
| 62c.. | Antenatal screening |
| 6776. | Pregnancy termination counselling |
| 679E. | Antenatal education |
| 7F051 | Diagnostic amniocentesis |
| 7F060 | Cerclage of cervix of gravid uterus |
| 7F25. | Obstetric monitoring |
| 7F2B1 | Ultrasound monitoring of early pregnancy |
| 8B75. | Vitamin supplement - pregnancy |
| 8H7W. | Refer to TOP counselling |
| 8HHV. | Referral for termination of pregnancy |
| 8HHf. | Refer to early pregnancy unit |
| 8HT9. | Referral to antenatal clinic |
| 8M6.. | Requests pregnancy termination |
| 95... | Maternity services admin. |
| 9N1N. | Seen in antenatal clinic |
| L1... | Pregnancy complications |
| L10.. | Haemorrhage in early pregnancy |
| L100. | Threatened abortion |
| L10y. | Other haemorrhage in early pregnancy |
| L11.. | Antepartum haemorrhage, abruptio placentae, placenta praevia |
| L1246 | Pre-eclampsia, unspecified |
| L13.. | Excessive pregnancy vomiting |
| L130. | Mild hyperemesis gravidarum |
| L1300 | Mild hyperemesis unspecified |
| L13y. | Other pregnancy vomiting |
| L13z. | Unspecified pregnancy vomiting |
| L1668 | Urinary tract infection complicating pregnancy |
| L16y5 | Abdominal pain in pregnancy |
| L1808 | Diabetes mellitus arising in pregnancy |
| L1809 | Gestational diabetes mellitus |
| L1825 | Iron deficiency anaemia of pregnancy |
| L18A0 | Cholestasis of pregnancy |
| L210. | Twin pregnancy |
| L25.. | Known or suspected fetal abnormality |

| | |
|---|---|
| L264. | Intrauterine death |
| L280. | Oligohydramnios |
| L33z. | Umbilical cord complications NOS |
| L413. | Antenatal deep vein thrombosis |
| L510. | Maternal care for hydrops fetalis |
| Lyu21 | [X]Other vomiting complicating pregnancy |
| Lyu25 | [X]Other specified pregnancy-related conditions |
| Z212. | Antenatal care |
| Z22.. | Pregnancy observations |
| Z225. | Normal pregnancy |
| Z226. | Pregnancy problem |
| Z227. | Confirmation of pregnancy |
| Z2291 | Intrauterine pregnancy |
| Z22A. | Observation of pattern of pregnancy |
| Z22A1 | Low risk pregnancy |
| Z22A4 | Early stage of pregnancy |
| Z22AA | Wanted pregnancy |
| Z22AB | Unplanned pregnancy |
| Z22B1 | Single pregnancy |
| Z22C1 | Estimated date of delivery from last period |
| Z22C3 | Length of gestation |
| Z22D1 | Viable pregnancy |
| Z22D2 | Non-viable pregnancy |
| Z22D3 | Uncertain viability of pregnancy |
| ZV222 | [V]Pregnancy confirmed |
| ZV223 | [V]Pregnant state, incidental |
| ZV231 | [V]Pregnancy with history of trophoblastic disease |
| ZV28. | [V]Antenatal screening |
| 1531. | Last menstural period-1st day |
| 271B. | O/E - fundal size = dates |
| 2726. | O/E - fetal presentation unsure |
| 2766. | O/E - fetal heart 120-160 |
| 5391. | Obstetric X-ray - fetus |
| 584A. | Dating/Booking US scan |
| 67A.. | Pregnancy advice |
| 67A2. | Diet in pregnancy advice |
| 67A3. | Pregnancy smoking advice |
| 67AE. | Folic acid advice in first trimester of pregnancy |
| 67AF. | Pregnancy advice for patients with epilepsy |
| 7F261 | Viability scan |
| 7F2B. | Obstetric ultrasound monitoring |
| 9511. | FP24 signed by patient |
| 957.. | FW 8-applic for presc exempt |

| | |
|---|---|
| 9kv.. | Pertussis vaccination programme pregnant women enhance service admin |
| 9mK.. | Pertussis vaccination in pregnancy invitation |
| 9Nk3. | Seen in fetal medicine clinic |
| 9NkN. | Seen in early pregnancy unit |
| 9NV1. | Antenatal clinic |
| ZV286 | [V] Antenatal screening for chromosomal anomalies |
| 6556. | Pretussis vaccination in pregnancy |

Supplementary table 2: ICD-10 version:2019 codes used to identify pregnancy from the hospital admissions data (PEDW)

| ICD-10 code | Definition |
|---|---|
| O11X | Pre-eclampsia superimposed on chronic hypertension |
| O120 | Gestational oedema |
| O121 | Gestational proteinuria |
| O122 | Gestational oedema with proteinuria |
| O13X | Gestational [pregnancy-induced] hypertension |
| O140 | Mild to moderate pre-eclampsia |
| O141 | Severe pre-eclampsia |
| O142 | HELLP syndrome |
| O149 | Pre-eclampsia, unspecified |
| O150 | Eclampsia in pregnancy |
| O116X | Unspecified maternal hypertension |
| O200 | Threatened abortion |
| O208 | Other haemorrhage in early pregnancy |
| O209 | Haemorrhage in early pregnancy, unspecified |
| O20 | Haemorrhage in early pregnancy |
| O210 | Mild hyperemesis gravidarum |
| O211 | Hyperemesis gravidarum with metabolic disturbance |
| O212 | Late vomiting of pregnancy |
| O219 | Vomiting of pregnancy, unspecified |
| O220 | Varicose veins of lower extremity in pregnancy |
| O223 | Deep phlebothrombosis in pregnancy |
| O224 | Haemorrhoids in pregnancy |
| O228 | Other venous complications in pregnancy |
| O229 | Venous complication in pregnancy, unspecified |
| O230 | Infections of kidney in pregnancy |
| O234 | Unspecified infection of urinary tract in pregnancy |
| O235 | Infections of the genital tract in pregnancy |
| O239 | Other and unspecified genitourinary tract infection in pregnancy |
| O244 | Diabetes mellitus arising in pregnancy |
| O249 | Diabetes mellitus in pregnancy, unspecified |

| | |
|---|---|
| O260 | Excessive weight gain in pregnancy |
| O261 | Low weight gain in pregnancy |
| O262 | Pregnancy care of habitual aborter |
| O265 | Maternal hypotension syndrome |
| O268 | Other specified pregnancy-related conditions |
| O269 | Pregnancy-related condition, unspecified |
| O280 | Abnormal haematological finding on antenatal screening of mother |
| O281 | Abnormal biochemical finding on antenatal screening of mother |
| O283 | Abnormal ultrasonic finding on antenatal screening of mother |
| O289 | Abnormal finding on antenatal screening of mother, unspecified |
| O300 | Twin pregnancy |
| O301 | Triplet pregnancy |
| O320 | Maternal care for unstable lie |
| O321 | Maternal care for breech presentation |
| O322 | Maternal care for transverse and oblique lie |
| O324 | Maternal care for high head at term |
| O326 | Maternal care for compound presentation |
| O328 | Maternal care for other malpresentation of fetus |
| O329 | Maternal care for malpresentation of fetus, unspecified |
| O340 | Maternal care for congenital malformation of uterus |
| O341 | Maternal care for tumour of corpus uteri |
| O342 | Maternal care due to uterine scar from previous surgery |
| O343 | Maternal care for cervical incompetence |
| O344 | Maternal care for other abnormalities of cervix |
| O346 | Maternal care for abnormality of vagina |
| O347 | Maternal care for abnormality of vulva and perineum |
| O348 | Maternal care for other abnormalities of pelvic organs |
| O350 | Maternal care for (suspected) central nervous system malformation in fetus |
| O351 | Maternal care for (suspected) chromosomal abnormality in fetus |
| O352 | Maternal care for (suspected) hereditary disease in fetus |
| O358 | Maternal care for other (suspected) fetal abnormality and damage |
| O359 | Maternal care for (suspected) fetal abnormality and damage, unspecified |
| O35 | Maternal care for known or suspected fetal abnormality and damage |
| O360 | Maternal care for rhesus isoimmunization |
| O361 | Maternal care for other isoimmunization |
| O363 | Maternal care for signs of fetal hypoxia |
| O364 | Maternal care for intrauterine death |
| O365 | Maternal care for poor fetal growth |
| O366 | Maternal care for excessive fetal growth |
| O368 | Maternal care for other specified fetal problems |
| O369 | Maternal care for fetal problem, unspecified |
| O40X | Polyhydramnios |
| O410 | Oligohydramnios |

| | |
|---|---|
| O418 | Other specified disorders of amniotic fluid and membranes |
| O429 | Premature rupture of membranes, unspecified |
| O438 | Other placental disorders |
| O440 | Placenta praevia specified as without haemorrhage |
| O441 | Placenta praevia with haemorrhage |
| O459 | Premature separation of placenta, unspecified |
| O468 | Other antepartum haemorrhage |
| O469 | Antepartum haemorrhage, unspecified |
| O470 | False labour before 37 completed weeks of gestation |
| O471 | False labour at or after 37 completed weeks of gestation |
| O479 | False labour, unspecified |
| O48X | Prolonged pregnancy |
| O718 | Other specified obstetric trauma |
| O882 | Obstetric blood-clot embolism |
| Z321 | Pregnancy confirmed |
| Z33X | Pregnant state, incidental |
| Z340 | Supervision of normal first pregnancy |
| Z348 | Supervision of other normal pregnancy |
| Z349 | Supervision of normal pregnancy, unspecified |
| Z352 | Supervision of pregnancy with other poor reproductive or obstetric history |
| Z353 | Supervision of pregnancy with history of insufficient antenatal care |
| Z357 | Supervision of high-risk pregnancy due to social problems |
| Z358 | Supervision of other high-risk pregnancies |
| Z368 | Other antenatal screening |
| Z369 | Antenatal screening, unspecified |

Supplementary Table 3. Rates and cumulative rates of vaccine uptake during pregnancy for all vaccinated women by month and age group

| Age | 18-24 | | | 25-29 | | | 30-39 | | | 40-50 | | | All age groups | | |
|---|---|---|---|---|---|---|---|---|---|---|---|---|---|---|---|
| | n | % | Cum % | n | % | Cum % | n | % | Cum % | n | % | Cum % | N | % | Cum % |
| Dec-20 | 22 | 2.0% | 0.5% | 51 | 2.3% | 0.7% | 75 | 1.7% | 0.6% | <5 | 1.2% | 0.5% | 152 | 1.9% | 0.6% |
| Jan-21 | 99 | 9.0% | 2.6% | 188 | 8.4% | 3.2% | 335 | 7.4% | 3.4% | 30 | 8.7% | 4.0% | 652 | 7.9% | 3.2% |
| Feb-21 | 48 | 4.4% | 3.6% | 86 | 3.9% | 4.4% | 131 | 2.9% | 4.5% | 11 | 3.2% | 5.3% | 276 | 3.4% | 4.3% |
| Mar-21 | 76 | 6.9% | 5.3% | 147 | 6.6% | 6.3% | 246 | 5.4% | 6.5% | 32 | 9.3% | 9.0% | 501 | 6.1% | 6.3% |
| Apr-21 | 34 | 3.1% | 6.0% | 158 | 7.1% | 8.5% | 585 | 12.9% | 11.3% | 75 | 21.7% | 17.7% | 852 | 10.4% | 9.7% |
| May-21 | 226 | 20.7% | 10.8% | 705 | 31.7% | 17.9% | 1469 | 32.4% | 23.4% | 74 | 21.4% | 26.4% | 2474 | 30.2% | 19.5% |
| Jun-21 | 253 | 23.1% | 16.3% | 225 | 10.1% | 20.9% | 444 | 9.8% | 27.1% | 44 | 12.8% | 31.5% | 966 | 11.8% | 23.4% |
| Jul-21 | 66 | 6.0% | 17.7% | 144 | 6.5% | 22.9% | 370 | 8.2% | 30.1% | 29 | 8.4% | 34.9% | 609 | 7.4% | 25.8% |
| Aug-21 | 98 | 9.0% | 19.8% | 217 | 9.8% | 25.8% | 401 | 8.8% | 33.4% | 20 | 5.8% | 37.2% | 736 | 9.0% | 28.7% |
| Sep-21 | 65 | 5.9% | 21.2% | 149 | 6.7% | 27.8% | 246 | 5.4% | 35.4% | 17 | 4.9% | 39.2% | 477 | 5.8% | 30.6% |
| Oct-21 | 41 | 3.7% | 22.0% | 55 | 2.5% | 28.5% | 104 | 2.3% | 36.3% | <5 | 0.9% | 39.6% | 203 | 2.5% | 31.5% |
| Nov-21 | 22 | 2.0% | 22.5% | 32 | 1.4% | 29.0% | 51 | 1.1% | 36.7% | <5 | 0.6% | 39.8% | 107 | 1.3% | 31.9% |
| Dec-21 | 44 | 4.0% | 23.5% | 68 | 3.1% | 29.9% | 82 | 1.8% | 37.4% | <5 | 1.2% | 40.3% | 198 | 2.4% | 32.7% |
| **Total** | **1094** | | | **2225** | | | **4539** | | | **345** | | | **8203** | | |

Supplementary Table 4. Vaccine uptake by Ethnicity/WIMD quintile of deprivation of area of residence and age group

| Age group | 18-24 | | | 25-29 | | | 30-39 | | | 40-50 | | |
|---|---|---|---|---|---|---|---|---|---|---|---|---|
| | Vaccinated | All | % | Vaccinated | All | % | Vaccinated | All | % | Vaccinated | All | % |
| Ethnicity | | | | | | | | | | | | |
| Asian | 28 | 101 | 27.7% | 92 | 269 | 34.2% | 191 | 492 | 38.8% | 20 | 40 | 50.0% |
| White | 887 | 3623 | 24.5% | 1763 | 5782 | 30.5% | 3685 | 9482 | 38.9% | 282 | 660 | 42.7% |
| Other | 23 | 92 | 25.0% | 52 | 153 | 34.0% | 108 | 296 | 36.5% | 11 | 30 | 36.7% |
| Mixed | 13 | 60 | 21.7% | 17 | 83 | 20.5% | 42 | 163 | 25.8% | <5 | 10 | 30.0% |
| Black | 7 | 54 | 13.0% | 29 | 128 | 22.7% | 61 | 226 | 27.0% | 8 | 32 | 25.0% |

| | | | | | | | | | | | | |
|---|---|---|---|---|---|---|---|---|---|---|---|---|
| Unknown | 136 | 734 | 18.5% | 268 | 1032 | 26.0% | 454 | 1482 | 30.6% | 23 | 87 | 26.4% |
| WIMD_Quintile_2019 | | | | | | | | | | | | |
| 5 (Least deprived) | 88 | 306 | 28.8% | 332 | 903 | 36.8% | 1024 | 2261 | 45.3% | 80 | 156 | 51.3% |
| 4th | 141 | 506 | 27.9% | 361 | 1110 | 32.5% | 834 | 2034 | 41.0% | 76 | 154 | 49.4% |
| 3rd | 197 | 733 | 26.9% | 367 | 1254 | 29.3% | 735 | 2034 | 36.1% | 42 | 136 | 30.9% |
| 2nd | 251 | 1071 | 23.4% | 447 | 1500 | 29.8% | 743 | 2093 | 35.5% | 53 | 131 | 40.5% |
| 1 (Most deprived) | 292 | 1582 | 18.5% | 458 | 1828 | 25.1% | 687 | 2257 | 30.4% | 58 | 173 | 33.5% |
| NA | 125 | 466 | 26.8% | 256 | 852 | 30.0% | 518 | 1462 | 35.4% | 38 | 109 | 34.9% |

Supplementary Table 5. Chi-square (*P* value) of COVID-19 vaccine uptake by pregnant women characteristics compared.

| Log Rank | X2 | P value | X2 | P value | X2 | P value | X2 | P value | X2 | P value | X2 | P value |
|---|---|---|---|---|---|---|---|---|---|---|---|---|
| Age | 18-24 | | 25-29 | | 30-39 | | 40-50 | | | | | |
| 18-24 | - | - | 0.46 | 0.50 | 30.32 | <.001 | 26.74 | <.001 | | | | |
| 25-29 | - | - | - | - | 42.73 | <.001 | 26.32 | <.001 | | | | |
| 30-39 | - | - | - | - | - | - | 4.75 | 0.029 | | | | |
| 40-50 | - | - | - | - | - | - | - | - | | | | |
| Ethnicity | Asian | | White | | Other | | Mixed | | Black | | Unknown | |
| Asian | - | - | 4.16 | 0.041 | 0.60 | 0.44 | 0.23 | 0.629 | 0.06 | 0.801 | 8.61 | 0.003 |
| White | - | - | - | - | 6.44 | 0.011 | 0.10 | 0.775 | 0.39 | 0.531 | 4.65 | 0.031 |
| Other | - | - | - | - | - | - | 0.76 | 0.385 | 0.42 | 0.519 | 11.17 | 0.001 |
| Mixed | - | - | - | - | - | - | - | - | 0.04 | 0.852 | 1.00 | 0.319 |
| Black | - | - | - | - | - | - | - | - | - | - | 1.97 | 0.161 |
| Unknown | - | - | - | - | - | - | - | - | - | - | - | - |
| WIMD | 1st | | 2nd | | 3rd | | 4th | | 5th | | Unknown | |
| 1st | - | - | 0.69 | 0.407 | 3.32 | 0.068 | 0.80 | 0.371 | 17.45 | <.001 | 3.45 | 0.063 |

| | | | | | | | | | | | | |
|---|---|---|---|---|---|---|---|---|---|---|---|---|
| 2nd | - | - | - | - | 6.80 | 0.009 | 0.05 | 0.823 | 12.90 | <.001 | 1.70 | 0.192 |
| 3rd | - | - | - | - | - | - | 7.24 | 0.007 | 36.13 | <.001 | 12.51 | <.001 |
| 4th | - | - | - | - | - | - | - | - | 12.24 | <.001 | 1.43 | 0.232 |
| 5th | - | - | - | - | - | - | - | - | - | - | 3.71 | 0.054 |
| Unknown | - | - | - | - | - | - | - | - | - | - | - | - |

### Supplementary Table 6. Demographics for women completing the survey

| | | **N** | **%** |
|---|---|---|---|
| **Age** | 18-24 | 21 | 6.3 |
| | 25-29 | 85 | 25.7 |
| | 30-39 | 147 | 44.4 |
| | 40-50 | 14 | 4.2 |
| | Unknown | 64 | 19.4 |
| **Ethnicity** | White | 272 | 82.2 |
| | Non-White | 5 | 1.5 |
| | Unknown | 54 | 16.3 |
